# Evaluating the Impact of a Self-Guided, Asynchronous, Balance Exercise Application on Fall-Related Injuries

**DOI:** 10.1101/2025.09.09.25335337

**Authors:** Kris F. Wain, Claudia A. Steiner, Andrea E. Daddato, Deanna B. McQuillan, John D. Litten, Chris Jentz, Andrew R. Jessen, Wendolyn S. Gozansky

## Abstract

**Background:** More than one in four older adults experience a fall each year. While exercise programs are effective in reducing fall-related injuries (FRI), participation remains low due to access barriers. The primary aim of this study was to evaluate whether older adults who registered for Nymbl, a self-guided, asynchronous, balance application, experienced fewer FRIs as compared to age-similar individuals who did not register.

**Methods:** This retrospective cohort study used data from Kaiser Permanente Colorado, linked to Nymbl registration and usage records based on patient name and demographic information between February 2018 and September 2024. The cohort included individuals aged 60 and older with continuous health plan enrollment for 12 months before and after Nymbl registration (or a randomly assigned index date). Logistic regression models estimated the association between Nymbl registration and FRIs during the 12-month follow-up, stratified by history of FRIs. Marginal effects reported the absolute risk difference associated with Nymbl registration. Secondary analyses examined dose-response effects of Nymbl usage and whether the effect of Nymbl was additive to participation in other exercise programs.

**Results:** We identified 3,735 individuals who registered for Nymbl and 114,219 age-eligible non-registrants. Among individuals with a prior FRI, Nymbl registration was associated with a 4.24 percentage point reduction in acute FRIs, however no significant effect was estimated for individuals without a baseline FRI. Secondary analysis indicated that at least five sessions were required to achieve a meaningful reduction in FRIs, and effects were limited to those not already participating in other exercise programs.

**Conclusion:** Findings from this study suggest that asynchronous, self-guided balance applications may reduce FRIs among older adults with a history of falls who are not otherwise engaged in structured exercise programs. Remotely delivered fall prevention programs may help overcome access barriers and can be used to supplement in-person and guided exercise programs.

## INTRODUCTION

More than one in four adults aged 65 and older experience a fall each year, resulting in approximately 3 million emergency department visits and 800,000 hospitalizations annually.^1^ In response to this significant healthcare burden, the U.S. Preventive Services Task Force (USPSTF) recommended exercise interventions to prevent falls in community-dwelling adults 65 years or older who are at increased risk with a Grade “B” recommendation, indicating moderate certainty of a net benefit.^2^ Despite the high prevalence of fall-related injuries (FRIs), participation in fall prevention programs remains low.^3,4^ Limited access remains a significant barrier, as many older adults struggle to attend in-person programs due to transportation challenges or scheduling constraints.^5,6^ To address access barriers, the USPSTF’s most recent recommendation emphasized the need for interventions that improve the availability and accessibility of effective fall prevention strategies, including asynchronous, remotely delivered exercise programs.^2^

Prior evaluations of asynchronous, remotely delivered fall prevention interventions are limited and primarily based on randomized trials. While findings have been mixed, access to a self-directed digital exercise program may reduce FRIs by up to 18%, particularly among individuals at low to intermediate fall risk.^7,8^ However, these studies have relied solely on patient-reported fall outcomes, which may be affected by recall bias, potentially leading to overreporting of minor events or underreporting of more serious fall injuries.^7,9^ Additionally, it remains unclear whether remotely delivered interventions offer benefits that are additive to those of traditional, in-person exercise programs. Moreover, individuals who participate in clinical trials tend to be healthier and more motivated than the broader population, which may limit the generalizability of trial findings.^10–12^

To better understand the effectiveness of asynchronous exercise programs in reducing FRIs using real-world evidence, the primary aim of this study was to evaluate whether community-dwelling older adults who registered for Nymbl, a self-guided balance training application, had a reduction in FRIs compared to age-similar adults who did not register. We examined the effect of Nymbl on both primary fall prevention (preventing initial FRIs) and secondary prevention (preventing recurrent FRIs). This research addresses a key knowledge gap identified by the USPSTF by evaluating the effectiveness of a digitally delivered fall prevention program, which supports efforts to expand access for individuals who face barriers to traditional in-person or guided exercise interventions.^2^

## METHODS

### Study Setting and Data

This study was conducted within Kaiser Permanente Colorado (KPCO), an integrated healthcare system providing both clinical care and insurance to approximately 550,000 members. KPCO’s coordinated care model delivers comprehensive care management for patients with FRIs, encompassing initial treatment, recovery-focused care, and outpatient follow-up.^13,14^ To improve physical well-being and help prevent FRIs, KPCO offers various exercise programs, such as Silver Sneakers®, a fitness program for older adults that provides free access to thousands of fitness centers and guided classes.

Through a partnership between Nymbl Science and the Denver Regional Council of Governments (DRCOG), all adults aged 60 and older living in the Denver Metro area were eligible to download the Nymbl app at no cost starting February 1, 2019. While available to anyone meeting the age and residence requirements, KPCO did not advertise or encourage Nymbl use among members during the study period. Nymbl is an asynchronous mobile application, accessible via smartphone or tablet, designed to improve balance and walking stability to help prevent falls.^15,16^ The Nymbl application uses a dual-tasking approach, combining physical activities such as balance or strength exercises with simultaneous cognitive challenges, like image recall. Evidence suggests that dual-task training can improve both static and dynamic balance, as well as executive function, particularly among older adults with a history of falls.^17^ A more detailed description of the Nymbl Balance application can be found in Supplemental Figure SF1.

Nymbl Science provided KPCO with a complete list of individuals who downloaded and registered for the Nymbl application during its partnership with DRCOG in accordance with a data transfer agreement. The list of Nymbl users was linked to KPCO members at the individual-level using name, birthdate, and demographic information. Data provided by Nymbl Science included the date of registration and timestamps of when the application was used. Electronic health record (EHR) and administrative data were obtained from the KPCO Virtual Data Warehouse, a standardized data repository containing information on diagnostic and procedural codes, patient-reported responses to health questionnaires, demographics, enrollment, and census based socioeconomic status measures.^18^ The cohort was limited to KPCO members aged 60+ who were insured between February 1, 2019 and September 30, 2024. Study protocol and human subjects considerations were reviewed and approved by the Kaiser Permanente Interregional Institutional Review Board (#2163506).

### Measures

The primary outcome was a FRI episode during the 365-day period after the index date. To quantify FRI episodes, we applied a previously published algorithm that identified FRIs using International Classification of Diseases version 10 diagnostic codes.^19^ This algorithm categorized FRI episodes into three levels of acuity based on injury type and care setting: (1) the **acute care algorithm**, which captured the most severe and clinically valid injuries, including those to the head, face, limbs, neck, and trunk treated in hospital or emergency department settings; (2) the **balanced algorithm**, which built upon the acute care definition by including severe injuries treated in non-emergency outpatient settings, offering a middle ground between specificity and comprehensiveness; and (3) the **inclusive algorithm**, which maximized FRI detection by incorporating nearly all injury types regardless of setting, allowing for broad identification of FRIs.

The primary exposure was registration for the Nymbl application. Participants were classified into two groups: individuals who registered for Nymbl, and an age-eligible comparison group who did not register for Nymbl. The date of registration was used as the index date among those who registered for Nymbl. Because the comparison group lacked a natural index date for evaluating FRI outcomes, each individual in the comparison group was assigned a randomly selected index date based on the distribution of registration dates among Nymbl users (Supplemental Figure SF2).^20,21^ This approach ensured equivalent follow-up intervals across both groups to assess FRI outcomes. All individuals were required to have continuous enrollment within KPCO for 12 months before and 12 months after their index date.

Baseline explanatory variables were measured during the 12-month period prior to the index date and included data on an individual’s demographics, comorbidity profile, healthcare utilization, and socioeconomic status. Age was grouped into three categories: 60–64 years, 65–75 years, and 76 years or older. Race was self-reported and categorized into mutually exclusive groups: Asian, Black, Hispanic, Unknown, White, and, due to small sample sizes, a combined single category labeled “Other Minority Races,” which included Native Hawaiian/Pacific Islander, Native American/Alaskan Native, and Multiracial individuals. Ethnicity was captured as a separate categorical variable indicating Hispanic, Non-Hispanic, or Unknown. Comorbidity burden was measured using the Deyo adaptation of the Charlson Comorbidity Index, based on diagnoses from the year prior to the index date.^22^ The number of diagnosed chronic conditions was grouped into four categories: 0 conditions, 1 condition, 2 conditions, and 3+ conditions. Utilization of acute medical services during baseline was captured using two binary indicators: one indicator for a hospital admission and a separate indicator for an emergency department visit.

Socioeconomic status was assessed using the Neighborhood Deprivation Index (NDI), a composite measure incorporating housing, poverty, employment, occupation, and education at the census tract level. Higher NDI values indicated greater levels of deprivation within the census tract.^23^

### Statistical Analysis

Baseline characteristics of Nymbl registrants and age-eligible non-registrants were compared using chi-square tests for categorical variables. Unadjusted differences in FRI outcomes between groups were calculated to help illustrate both absolute differences and relative changes in FRI outcomes. Logistic regression models were used to evaluate the association of Nymbl registration on FRI outcomes during the 12 months after the index date. To better understand Nymbl’s association with both primary fall prevention (preventing initial FRI) and secondary prevention (preventing recurrent FRI), our primary analysis stratified individuals by whether they experienced any FRI in the 12-month period prior to the index date. Separate models were estimated for each of the three FRI acuity outcomes: acute, balanced, and inclusive, to determine whether Nymbl’s impact varied by severity of the FRI episode. Marginal effects (ME) and 95% confidence intervals (CI) were reported holding all other explanatory variables at their mean values. In this study, the ME is interpreted as the percentage point (pp) difference in the probability of an FRI outcome between Nymbl registrants and the age-eligible comparison group.

### Secondary and Sensitivity Analysis

To assess whether the amount of Nymbl utilization was associated with differential effects on FRI outcomes, we conducted a dose-response analysis limited to individuals who registered for Nymbl. We summed the number of Nymbl sessions for each individual during the first 90 days after registration and categorized them into three groups: 0 sessions (registered but did not use Nymbl), 1–4 sessions, >=5 sessions. FRI outcomes were then assessed during days 91 to 365 post-registration. Because we allowed for three levels of exposure, multinomial regression models were used to estimate the dose-response effect using registrants with 0-sessions as the referent group. Due to a sharp reduction in sample size when limiting our cohort to Nymbl users, it was necessary to collapse race into White vs. Non-White and diagnosed chronic conditions were regrouped as 0, 1–2, or 3+ conditions to maintain model stability.

In a complementary analysis, we used data from a health risk assessment (HRA) for Medicare beneficiaries as an additional independent variable to determine whether patient-reported fall risk factors yielded estimates similar to those based on pre-index FRI episodes identified using EHR data. The HRA collects self-reported medical, functional, and social data, including information on recent falls and balance concerns (Supplemental Figure SF3).^24^ Among individuals in our cohort who completed the HRA, we re-estimated logistic regression models stratified by a binary variable indicating self-reported fall or balance impairment.

To evaluate whether the effects of Nymbl were additive to other exercise interventions which may reduce FRIs, we re-estimated our primary regression models further stratifying by whether individuals participated in the Silver Sneakers® fitness program at any point during the 12-month pre-index period. All analyses were performed using SAS® Software version 9.4 (SAS Institute Inc., Cary, North Carolina) and STATA® Statistical Software version 18.0 (StataCorp. 2019. Stata Statistical Software: Release 16. College Station, TX: StataCorp LLC).

## RESULTS

After applying exclusion criteria, our cohort included 117,954 individuals (Supplemental Figure SF4), 3,735 (3.2%) who registered for Nymbl, and 114,219 (96.8%) who were age-eligible but did not register for Nymbl (Table 1). Compared to age-eligible non-registrants, Nymbl registrants were more likely to have a pre-index FRI (*Acute*: 3.1% vs. 2.0%, *Balanced*: 3.4% vs 2.2%, *Inclusive*: 5.2% vs 3.1%), be 76 or older (35.6% vs. 28.1%), female (72.8% vs. 54.9%), White (90.1% vs. 77.1%), and reside in the least deprived census tracts (48.8% vs. 42.0%). No statistically significant differences were observed between groups in the number of diagnosed chronic conditions or in utilization of acute medical services during the pre-index period.

**Table 1.** Baseline Characteristics by Nymbl Registration.

|  | **Nymbl User** |  |  |  |
| --- | --- | --- | --- | --- |
|  | *No* | *Yes* | *Total* | *p-value* |
| **Unique Patients, N (%)** | 114,219 (96.8) | 3,735 (3.2) | 117,954 |  |
| **Pre-Index Fall-Related Injury, N (%)** |  |  |  |  |
| Acute Algorithm | 2,302 (2.0) | 117 (3.1) | 2,419 (2.1) | *< .0001* |
| Balanced Algorithm | 2,541 (2.2) | 128 (3.4) | 2,669 (2.3) | *< .0001* |
| Inclusive Algorithm | 3,594 (3.1) | 194 (5.2) | 3,788 (3.2) | *< .0001* |
| **Age at Index Date, N (%)** |  |  |  | *< .0001* |
| 60 to 64 | 25,830 (22.6) | 417 (11.2) | 26,247 (22.3) |  |
| 65 to 75 | 56,344 (49.3) | 1,990 (53.3) | 58,334 (49.5) |  |
| 76 and older | 32,045 (28.1) | 1,328 (35.6) | 33,373 (28.3) |  |
| **Gender, N (%)** |  |  |  | *< .0001* |
| Female | 62,728 (54.9) | 2,718 (72.8) | 65,446 (55.5) |  |
| **Race, N (%)** |  |  |  | *< .0001* |
| Asian | 3,375 (3.0) | 31 (0.8) | 3,406 (2.9) |  |
| Black | 4,704 (4.1) | 72 (1.9) | 4,776 (4.0) |  |
| White | 88,069 (77.1) | 3,367 (90.1) | 91,436 (77.5) |  |
| Native Hawaiian / Pacific Islander,<br>American Indian / Alaska Native,<br>Multi-Race, or Other Race | 14,907 (13.1) | 217 (5.8) | 15,124 (12.8) |  |
| Unknown | 3,164 (2.8) | 48 (1.3) | 3,212 (2.7) |  |
| **Hispanic, N (%)** |  |  |  | *< .0001* |
| Yes | 11,742 (10.3) | 133 (3.6) | 11,875 (10.1) |  |
| No | 98,999 (86.7) | 3,531 (94.5) | 102,530 (86.9) |  |
| Missing | 3,478 (3.0) | 71 (1.9) | 3,549 (3.0) |  |
| **Charlson Comorbidity Index, N (%)** |  |  |  | *0.8822* |
| 0 | 81,324 (71.2) | 2,672 (71.5) | 83,996 (71.2) |  |
| 1 | 11,148 (9.8) | 369 (9.9) | 11,517 (9.8) |  |
| 2 | 8,873 (7.8) | 278 (7.4) | 9,151 (7.8) |  |
| 3 or more | 12,874 (11.3) | 416 (11.1) | 13,290 (11.3) |  |
| **Utilization (binary indicator), N (%)** |  |  |  |  |
| Emergency Department | 17,237 (15.1) | 6,04 (16.2) | 17,841 (15.1) | *0.0698* |
| Inpatient Admission | 8,656 (7.6) | 316 (8.5) | 8,972 (7.6) | *0.0454* |

| Neighborhood Deprivation Index Quintile, N (%) |  |  |  | < .0001 |
| --- | --- | --- | --- | --- |
| 1 (least deprived) | 47,981 (42.0) | 1,821 (48.8) | 49,802 (42.2) |  |
| 2 | 30,352 (26.6) | 1,031 (27.6) | 31,383 (26.6) |  |
| 3 | 17,560 (15.4) | 498 (13.3) | 18,058 (15.3) |  |
| 4 | 14,449 (12.7) | 312 (8.4) | 14,761 (12.5) |  |
| 5 (most deprived) | 3,877 (3.4) | 73 (2.0) | 3,950 (3.3) |  |

– *Neighborhood Deprivation Index measures 13 dimensions of socioeconomic status at the census-tract level including: wealth and income, education, occupation, and housing conditions*

Unadjusted analyses are shown in Supplemental Table ST1. Among individuals with a pre-index FRI, those who registered for Nymbl were 4.90pp less likely to experience an acute FRI (3.61% vs. 8.51%), 4.43pp less likely to experience a balanced FRI (4.64% vs. 9.07%), and 3.91pp less likely to experience an inclusive FRI (7.22% vs. 11.13%) compared to non-Nymbl users.

Primary results from logistic regression models stratified by pre-index FRI status are presented in Figure 1. Among individuals with a pre-index FRI episode, Nymbl registration was associated with a 4.24pp reduction in acute FRIs (ME = –4.24; 95% CI: –6.75 to –1.73), a 3.82pp reduction in balanced FRIs (ME = –3.82; 95% CI: –6.66 to -0.98), and a 3.70pp reduction in inclusive FRIs (ME = –3.70; 95% CI: –7.30 to –0.11). In contrast, among individuals who did not have a pre-index FRI, no significant association was estimated between Nymbl registration and FRI outcomes.

**Figure 1.**
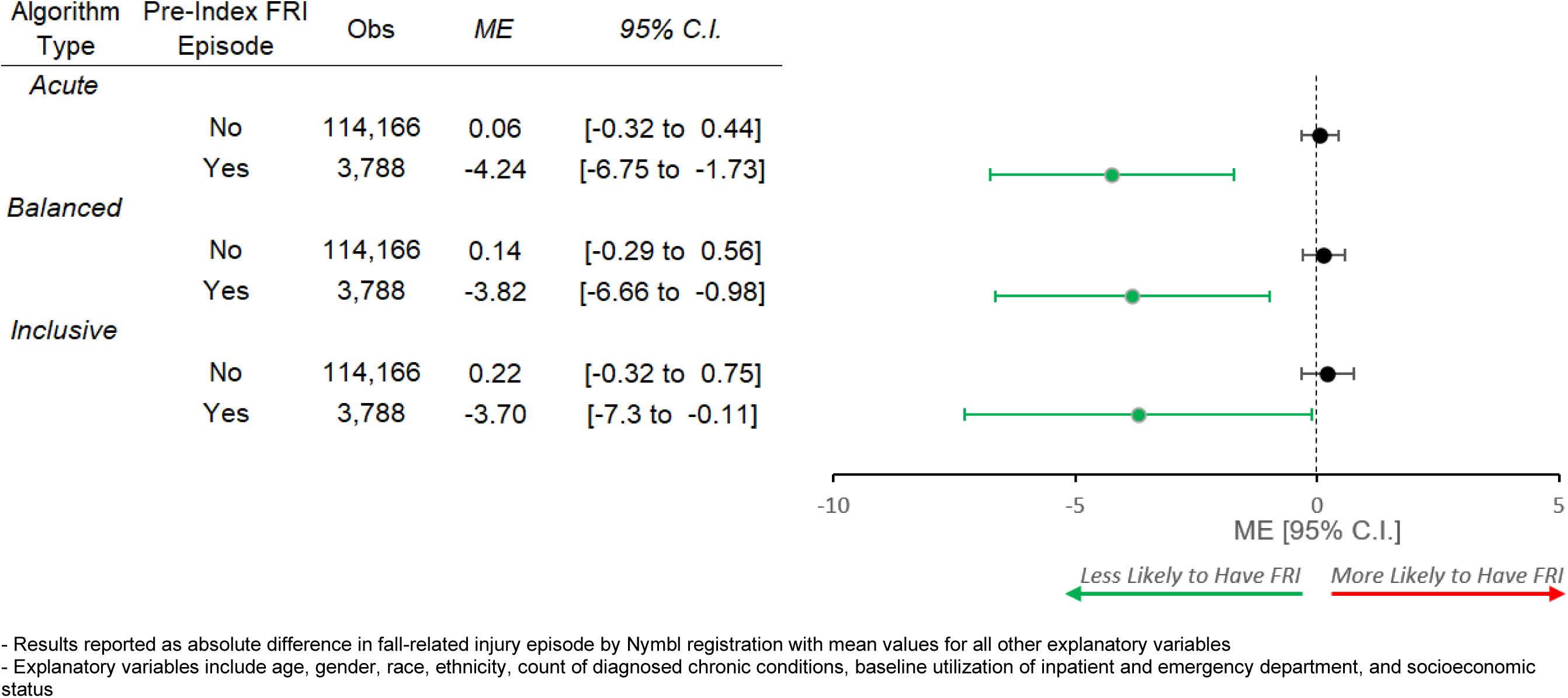
Adjusted Marginal Effect (ME) of Nymbl Registration on Occurrence of a Fall-Related Injury (FRI) Episode by Acuity Level Stratified by Pre-Index FRI Episode

Estimates from dose-response analyses among Nymbl users are shown in Figure 2. Compared to individuals who registered but did not use Nymbl, logging five or more sessions was associated with a 0.78pp reduction in acute FRIs (ME = –0.78; 95% CI: –1.49 to –0.07), a 1.15pp reduction in balanced FRIs (ME = –1.15; 95% CI: –2.14 to –0.16), and a 1.61pp reduction in inclusive FRIs (ME = –1.61; 95% CI: –2.95 to –0.27). No significant associations were estimated among individuals who used Nymbl 1 to 4 times compared those who registered but did not use Nymbl.

**Figure 2.**
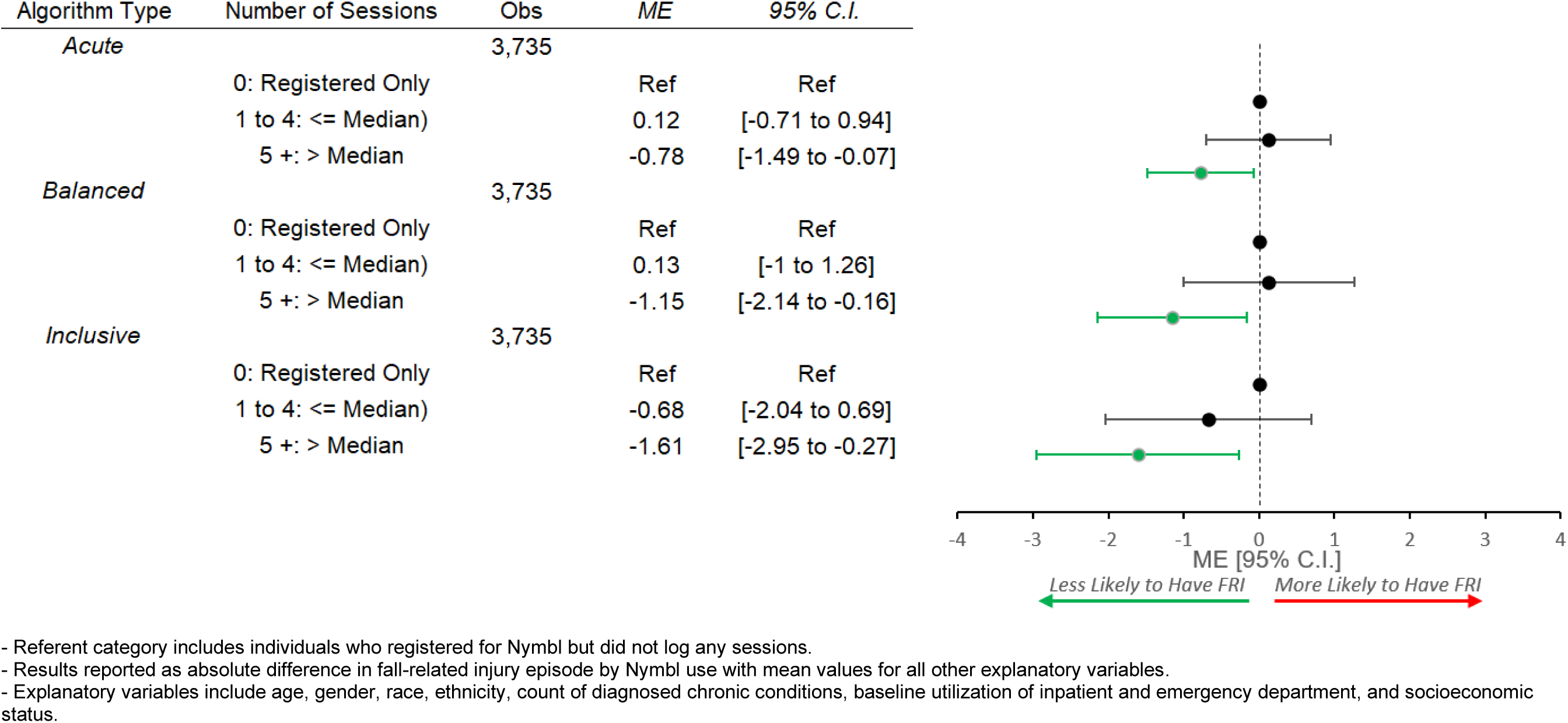
Among Nymbl Registrants: Adjusted Marginal Effect (ME) of Nymbl Use During 90 Days Post-Registration on FRI Episodes During Days 91 to 365 Post-Registration (Dose-Response)

Results using self-reported fall or balance impairment as the primary exposure variable are shown in Figure 3. Among individuals who reported a recent fall or difficulty with balance, Nymbl use was associated with a non-significant 1.29pp reduction in acute FRIs (ME = –1.29; 95% CI: –2.70 to 0.11), and no significant reductions estimated for balanced or inclusive FRIs in this group. Additionally, Nymbl use was not associated with a reduction in FRI episodes at any acuity level among individuals who did not report fall or balance concerns.

**Figure 3.**
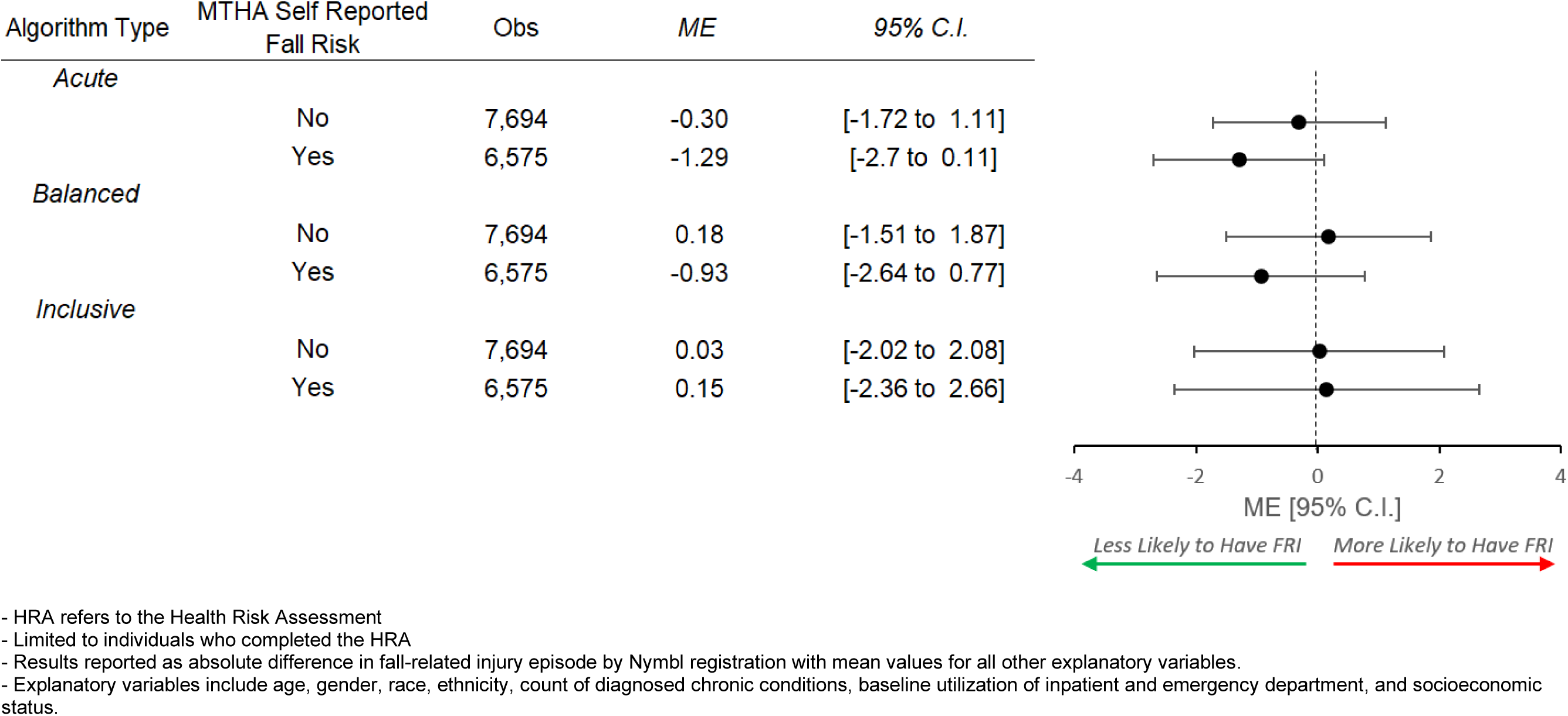
Adjusted Marginal Effect (ME) of Nymbl Registration on Occurrence of a Fall-Related Injury Episode by Acuity Level Stratified by Self-Reported Risk of Fall or Balance Difficulty

Estimates stratified by both pre-index FRI episode and participation in the Silver Sneakers® fitness program are shown in Figure 4. Among individuals who had a pre-index FRI but did not participate in Silver Sneakers®, we estimated that Nymbl was associated with a 4.93pp reduction in acute FRIs (ME = –4.93; 95% CI: –7.75 to –2.11), a 4.75pp reduction in balanced FRIs (ME = –4.75; 95% CI: –7.93 to –1.57), and a 5.73pp reduction in inclusive FRIs (ME = –5.73; 95% CI: –9.56 to –1.90). No significant associations were estimated for individuals with a pre-index FRI episode who participated in Silver Sneakers®, or for those without a pre-index FRI episode, regardless of Silver Sneakers® participation.

**Figure 4.**
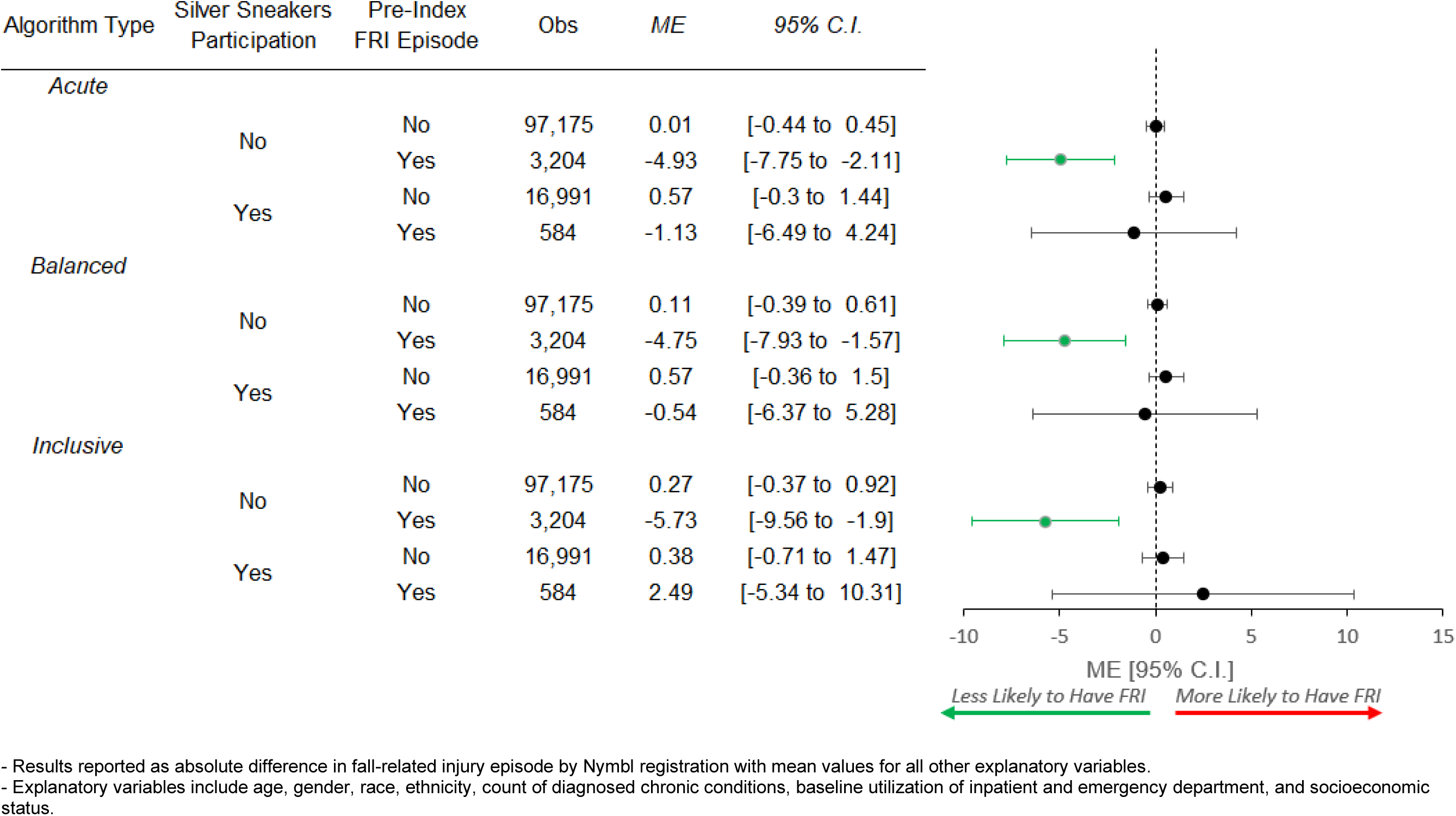
Adjusted Marginal Effect (ME) of Nymbl Registration on Occurrence of a Fall-Related Injury (FRI) Episode by Acuity Level Stratified by Pre-Index FRI Episode and Silver Sneakers Participation

## DISCUSSION

In this study, the use of an asynchronous, digital application designed to improve balance was associated with a 4.24pp reduction in acute FRI episodes (equivalent to a 57% relative decrease) among individuals with any prior FRI (acute, balanced, or inclusive). These findings suggest that self-directed digital fall prevention programs may provide an effective and more accessible alternative to in-person or instructor-led exercise programs.^25^ In comparison, findings from prior randomized trials of self-directed, unsupervised digital exercise programs have reported fall injury reductions between 8% to 18% among at risk populations.^7,8^ However, these trials relied on self-reported questionnaire data, which may have resulted in overreporting of minor fall injuries and consequently underestimated the impact of the intervention on acute fall injuries.

The cohort in the present study and in both of the prior randomized trials were predominantly White females, underscoring the need for targeted efforts to increase participation among minority and male populations. While fall risk may vary by sex, race, and ethnicity, fall related mortality rises with age across all groups and has been increasing over the last 20 years.^26^ Personalizing digital fall-reduction interventions to align with individual preferences and cultural contexts may improve engagement and increase accessibility across diverse populations.^5^

Findings from our dose-response analysis among Nymbl registrants indicated that completing at least five sessions was necessary to reduce FRIs. However, of the 3,735 individuals who registered for Nymbl, only 789 (21%) completed five or more sessions. This is consistent with previous studies showing that most participants in fall prevention interventions do not achieve the recommended dose of exercise activity.^7,8^ Given the abundance of digital exercise applications, sustaining user engagement over longer time horizons is critical to realizing the full benefits of these programs. As digital fall prevention strategies become increasingly embedded into routine care, efforts should focus on promoting sustained engagement through approaches such as integration with primary care, personalized goal setting, and continuous refinement of the user experience.^27–29^

Analysis using self-reported fall risk factors as the primary exposure variable found that Nymbl was associated with non-significant, but potentially clinically meaningful reductions in acute FRI episodes. Prior studies have shown balance and gait impairment to be a key predictor of more severe FRIs among community-dwelling older adults.^30^ However, relying on patient reported risk factors may be insufficient to adequately identify individuals at high risk of FRIs. One study found that fewer than 40% of healthcare providers routinely asked older adults about their fall history, highlighting significant missed opportunities for prevention.^3^ Additionally, fall-related care is often reactive, initiated after a fall has occurred, rather than preventive, even among high-risk individuals.^4^ While gait speed has been shown as a risk factor for FRIs, the association between gait speed and fall risk is non-linear. Individuals with faster gait speeds may be more agile and experience less severe injuries, while slower gait speeds may reflect underlying frailty, increasing the likelihood of more serious FRIs, complicating efforts to predict FRIs.^31,32^ Moreover, long-term risk factors such as impaired balance and gait speed can interact with short-term triggers, including acute illness or environmental hazards, adding additional complexity to FRI prediction.^33–35^ Although many fall risk assessment tools are available, current evidence suggests that no single measure offers sufficient accuracy to predict FRIs in community-dwelling older adults.^36,37^ As such, a patient’s history of prior falls remains one of the most reliable indicators of future FRI risk.^36^

Sensitivity analyses evaluating the impact of Nymbl among individuals who were already participating in an exercise program indicated that Nymbl’s effect was limited to those who were not taking part in the Silver Sneakers® program. This finding suggests that engaging in exercise, regardless of its primary intent or mode of delivery, may be effective in reducing FRIs. Prior evidence has suggested that brisk walking can be more effective than structured balance training in preventing fall injuries.^38^ Offering individuals a choice in program format (e.g., in-person vs. virtual, synchronous vs. asynchronous, guided vs. self-directed) may enhance long-term engagement, contributing to larger and more sustained reductions in FRI episodes.

## LIMITATIONS

This study has several limitations. Participation in Nymbl was voluntary. If those who chose to register for Nymbl were more aware of their fall risk, motivated, or simultaneously engaged in other fall prevention strategies, the observed effect of Nymbl may be overstated. Because the study cohort was predominantly White, non-Hispanic females, our findings may not extend to more diverse populations. While our EHR data likely captured hospitalizations and emergency department visits related to severe fall injuries, less severe FRIs may have gone unreported for individuals who did not seek medical care.^39,40^ If individuals who registered for Nymbl were more engaged with the healthcare system, they may have had more opportunities to report FRIs and been less susceptible to recall bias than non-registrants, potentially resulting in an underestimation of Nymbl’s true impact.^33^ Additionally, the follow-up period was limited to 12 months after the index date; thus, we could not assess the long-run effects of Nymbl on FRI episodes.

## CONCLUSION

Findings from this study suggest that asynchronous, self-directed digital balance programs may reduce FRIs, strengthening the evidence for integrating remote digital interventions into USPSTF guidelines for fall prevention.^2^ However, the effectiveness of these applications may be limited to individuals with a history of prior FRIs and those not already engaged in other exercise programs, thus implementation should target high-risk, sedentary populations. With the increasing number of fall prevention applications readily available, sustained patient engagement will be essential to realizing the full impact of digitally delivered interventions. Future research can examine the long-term effects of asynchronous exercise programs and explore their potential to generate cost savings and injury-prevention for both healthcare systems and patients.

## Data Availability

The data that support the findings of this study cannot be made publicly available because it contains sensitive patient information and is subject to strict privacy regulations under HIPAA

## SUPPLEMENTAL MATERIALS

**Supplemental Figure SF1.**
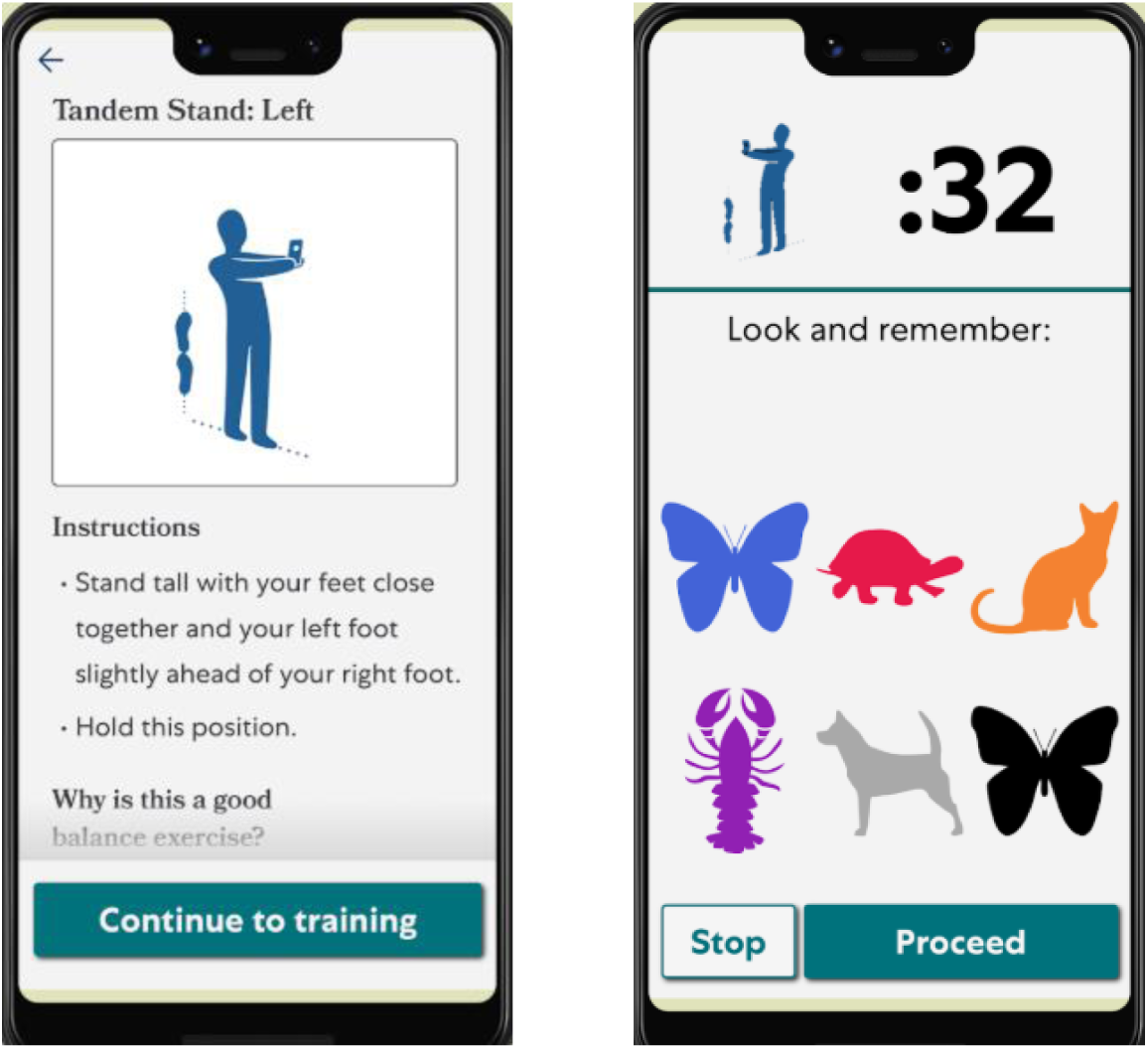
Description of Nymbl Balance Application

The Nymbl app is a digital platform specifically designed to deliver exercise interventions to older adults. It employs an innovative technique known as dual-task training, which combines physical exercises, such as balance or strength activities, with simultaneous cognitive challenges, like memory recall tasks. This approach targets an individual’s dual-task ability, or the capacity to perform two tasks at once, which tends to decline with age due to physical deconditioning and cognitive decline. Diminished dual-task ability can result in slower reaction times and an increased risk of falls.

Research has shown that dual-task training can improve both static and dynamic balance as well as executive functioning in older adults with a history of falls. In addition to dual-task training, Nymbl incorporates cognitive behavioral training to address psychological contributors to fall risk, such as fear of falling, which can itself limit physical activity and mobility.

**Supplemental Figure SF2.**
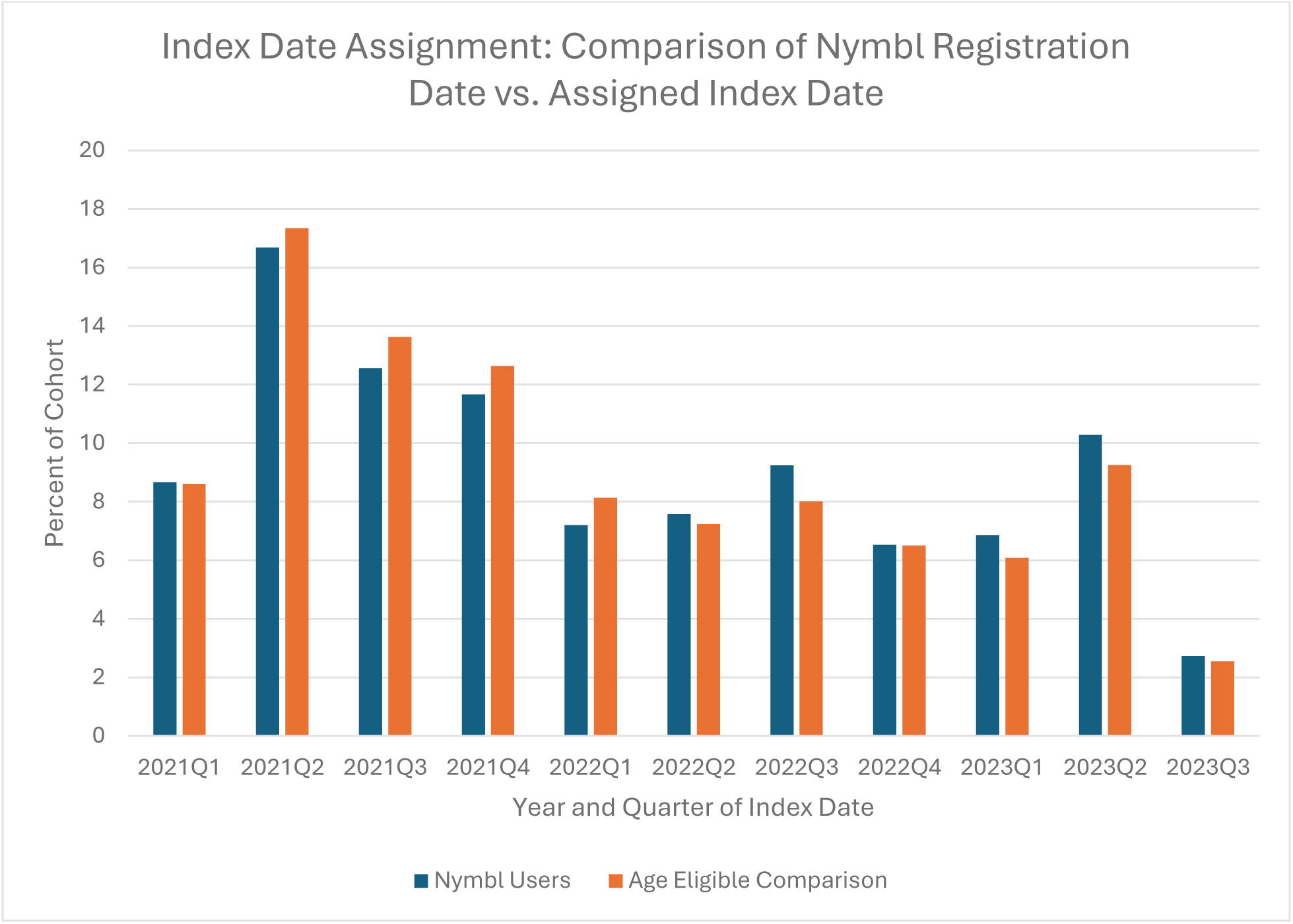
Distribution of Index Dates Among Nymbl Participants as Compared to Distribution of Randomly Assigned Index for Age-Eligible Non-Participants

For individuals who registered for Nymbl, the date of registration served as the index date. In contrast, members of the age-eligible comparison group who did not register lacked a natural reference point to anchor fall-related injury (FRI) outcomes. To enable valid comparisons between groups, a random index date was assigned to non-registrants using a method adapted from Jacob et al.^20^ Specifically, for each Nymbl registrant, we calculated the number of days between the study start date (February 1, 2019) and their registration date. These intervals were then pooled across all users to establish a distribution of index date probabilities. Using this distribution, index dates were randomly assigned to individuals in the comparison group to mirror the temporal distribution of Nymbl users. FRI outcomes were assessed over the 12-month period following each individual’s index date. The figure above displays the distribution of index dates for both Nymbl registrants and the comparison group, aggregated by year and quarter.

**Supplemental Figure SF3.**
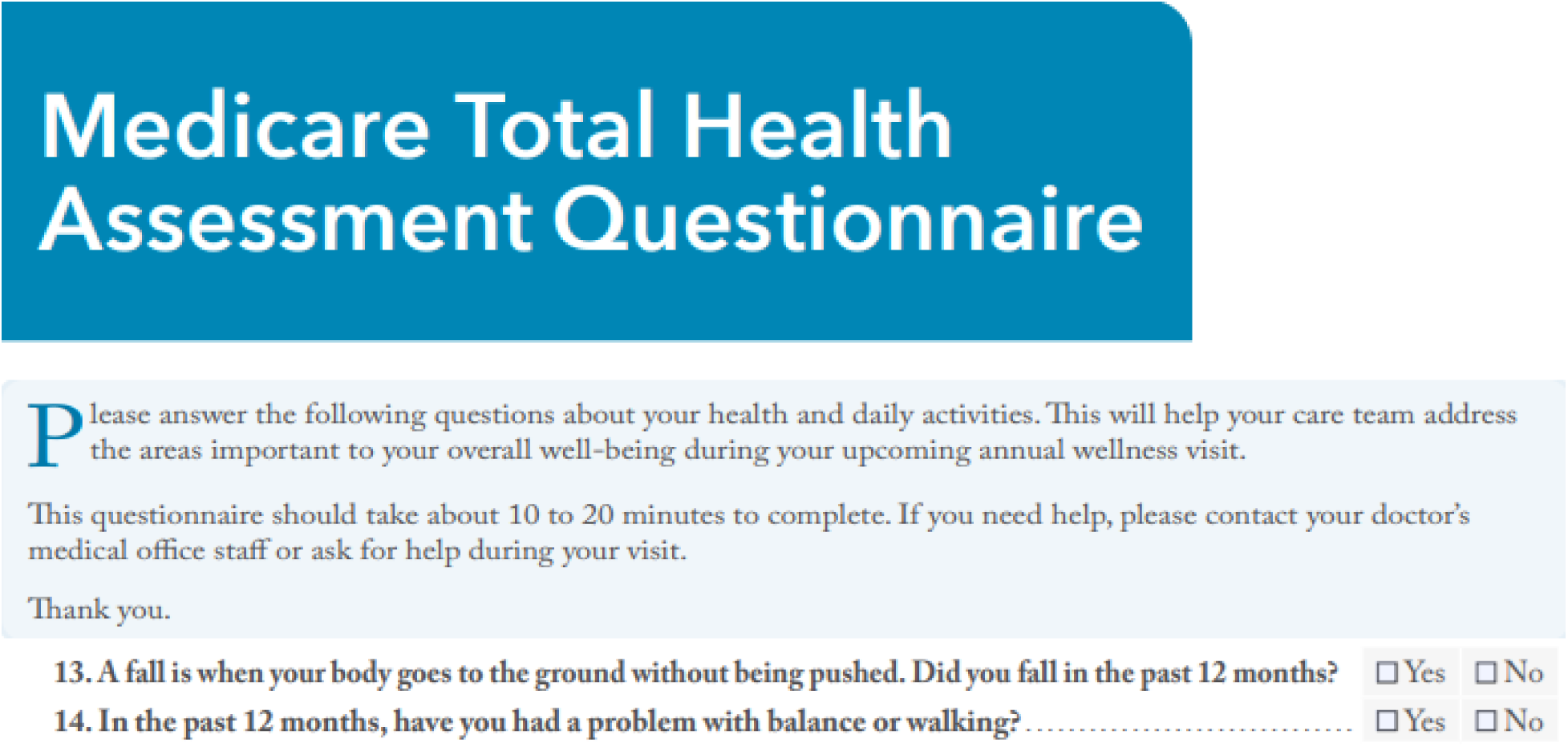
Patient Self-Reported Fall-Related Injury Risk Factors from the health risk assessment (HRA) tool

Two questions on the HRA specifically address prior falls and balance concerns as shown in Figure SF2. Patients who responded “yes” to either question were classified as having reported a fall-related injury (FRI) risk factor. Due to small sample size, responses to these two items were combined into a single binary variable indicating the presence of any self-reported FRI risk.

**Supplemental Figure SF4.**
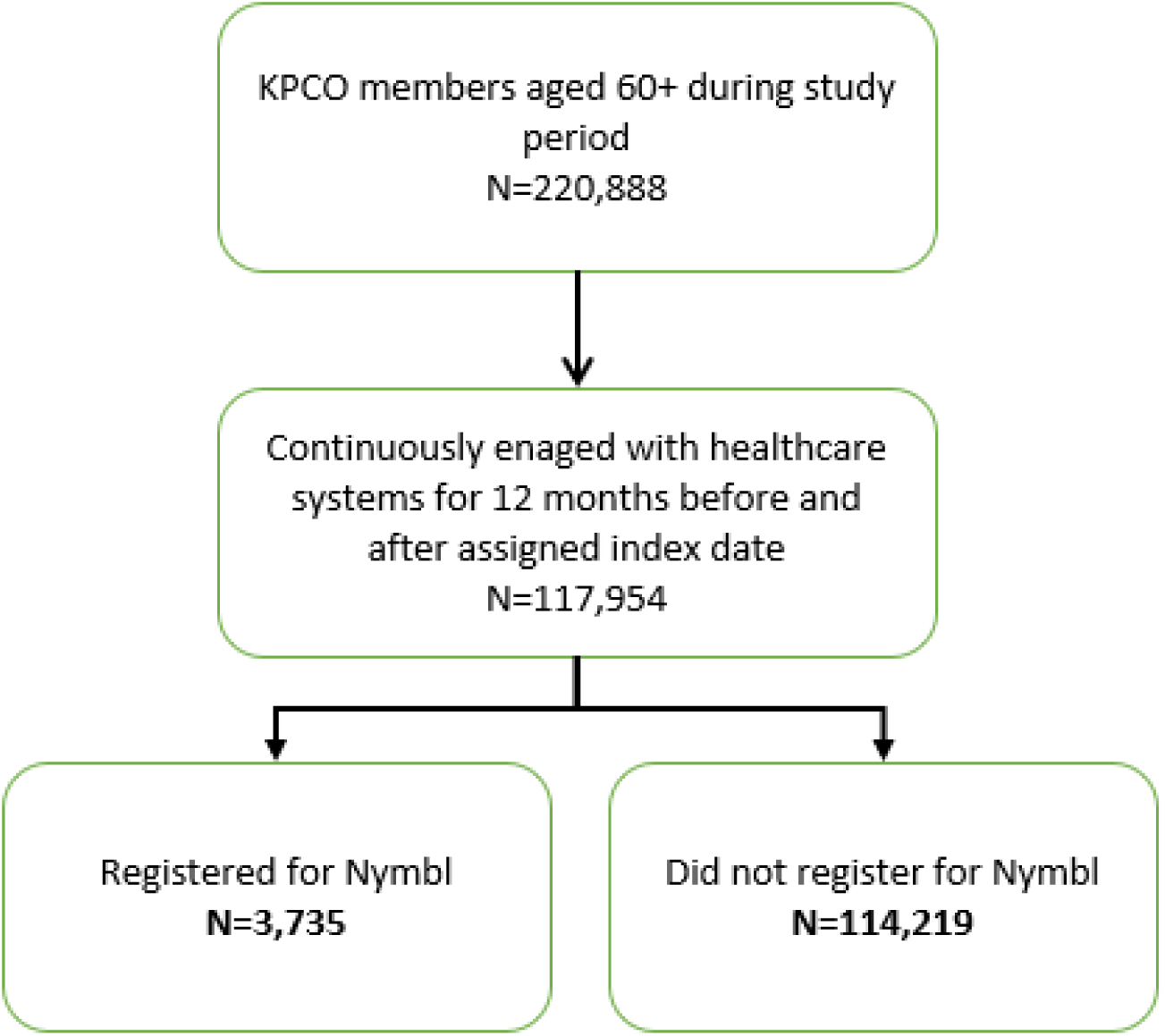
Consort Diagram

**Supplemental Table ST1.**
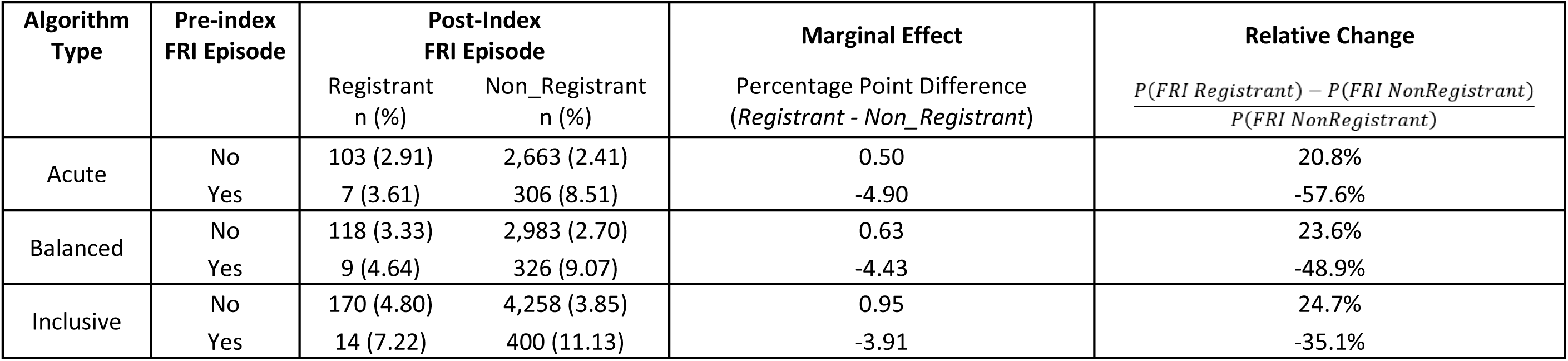
Unadjusted Marginal Effect (ME) of Nymbl Registration on Occurrence of a Fall-Related Injury (FRI) Episode by Acuity Level Stratified by Pre-Index FRI Episode.

